# qPCR in a suitcase for rapid *Plasmodium falciparum* and *Plasmodium vivax* surveillance in Ethiopia

**DOI:** 10.1101/2022.04.13.22273827

**Authors:** Lise Carlier, Sarah Cate Baker, Tiffany Huwe, Delenasaw Yewhalaw, Werissaw Haileselassie, Cristian Koepfli

## Abstract

Many *Plasmodium* spp. infections, both in clinical and asymptomatic patients, are below the limit of detection of light microscopy or rapid diagnostic test (RDT). Molecular diagnosis by qPCR can be valuable for surveillance, but is often hampered by absence of laboratory capacity in endemic countries. To overcome this limitation, we optimized and tested a mobile qPCR laboratory for molecular diagnosis in Ziway, Ethiopia, where transmission intensity is low. Protocols were optimized to achieve high throughput and minimize costs and weight for easy transport. 899 samples from febrile patients and 1021 samples from asymptomatic individuals were screened by local microscopy, RDT, and qPCR within a period of six weeks. 34/52 clinical *Plasmodium falciparum* infections were missed by microscopy and RDT. Only 4 asymptomatic infections were detected. No *hrp2* deletions were observed among 25 samples typed, but 19/24 samples carried *hrp3* deletions. The majority (25/41) of *Plasmodium vivax* infections (1371 samples screened) were found among asymptomatic individuals. All asymptomatic *P. vivax* infections were negative by microscopy and RDT. In conclusion, the mobile laboratory described here can identify hidden parasite reservoirs within a short period of time, and thus inform malaria control activities.

## Introduction

Malaria remains a major public health threat in many countries in the tropics and subtropics. After a decade of progress with a pronounced reduction of the number of clinical cases and deaths, progress has stalled in recent years. In 2020, over 240 million cases and 600,000 deaths were recorded [1].

Accurate and fast diagnosis and treatment are key aspects of malaria control. In most malaria-endemic countries, diagnosis by light microscopy is routinely conducted at health centers and hospitals. The sensitivity and specificity of local microscopy depends greatly on the training of local microscopists [2], and field microscopy can be substantially less sensitive than expert microscopy [3]. As an alternative, rapid diagnostic tests (RDTs) have become increasingly common. RDTs are lateral flow devices that detect parasite-specific proteins through immunohistochemistry. RDTs require less training, and results are obtained within 10 minutes. They are thus used by small health posts with no microscopy infrastructure and by health workers conducting household visits and diagnosis, e.g. in the frame of reactive case detection activities [4]. Sensitivity of RDTs can be impaired by incorrect storage and handling, wrong interpretation of results, or deletion of the gene coding for Histidine-Rich Protein 2 (HRP2), which is detected by most RDTs for *P. falciparum* [5]. False-positive results can be caused by non-malarial infections [6].

Light microscopy and RDT have a limit of detection of approximately 50-100 parasites per uL of blood [7]. A large number of clinical infections remain below this density [8, 9]. Further, in all transmission settings, a proportion of infections remain asymptomatic, and many of them are subpatent [10, 11]. Asymptomatic infections and low-density clinical infections escaping routine diagnosis conducted at health centers among febrile patients sustain transmission and present a major challenge to control [12-15].

Molecular diagnosis by PCR or other nucleic acid amplification tests are required to assess the quality of local diagnosis, to determine the true number of infections among febrile patients, and to understand population parasite prevalence in asymptomatic individuals. Rapid, sensitive screening might also be required to coordinate the response to outbreaks, for example to decide where intensified vector control is warranted because of a large asymptomatic reservoir. Molecular surveillance is often complicated by the absence of laboratory infrastructure and lack of skilled personnel in malaria endemic sites. Shipment of samples to reference laboratories can be complicated and time consuming. Molecular screening is thus seldom applied to select control strategies tailored to local conditions, or in response to outbreaks.

In order to speed up time to result and enable in-country scientists and control programs to process samples, efforts are increasingly being made to bring laboratory capacity to field sites [16-18]. Numerous devices and protocols for molecular screening for pathogens are being developed and trialed. Often, these assays rely on custom-built devices [19]. Throughput of commercially available platforms is often low [20-24]. In addition, the need for high-throughput, mobile DNA extraction platforms is not addressed.

For this study, a mobile qPCR lab was trialed for malaria surveillance in a low transmission site in Ethiopia. All equipment and consumables needed are commercially available and fit in suitcases for transport on airplanes. Up to two 96-well plates can be processed in a day, at a cost of approximately USD 2.5 per sample for DNA extraction and *P. falciparum* and *P. vivax* qPCR. Within a brief period of 2 months, nearly 2000 samples from febrile cases and asymptomatic individuals were screened using highly sensitive qPCR.

## Methods

### Ethical approval

Informed written consent was obtained prior to sample collection from each study participant or, in the case of minors, from their parent or legal guardian. The study protocol was approved by the University of Notre Dame IRB (#19-03-5201), Trinity College Dublin, Addis Ababa University, and the National Research Ethics Review Committee at Ministry of Science and Higher education (MoSHE).

### Study site

In Ethiopia, *P. falciparum* and *P. vivax* are endemic. Malaria transmission ranges from very high in the tropical lowlands along the borders with Sudan and South Sudan to low and sporadic in the highlands [25]. In 2019, over 900,000 confirmed cases were reported. This represents a pronounced reduction compared to 2013, where the number of cases peaked at 2.6 million, but only a moderate reduction compared to 2010, with 1.2 million confirmed cases [26]. Diagnosis is provided at over 20,000 health centers across the country. Larger health centers diagnosis by microscopy, while RDTs are used by smaller health posts. In addition, over 70,000 health extension workers visit households and provide basic medical services, including malaria diagnosis by RDT [27].

This study was conducted in Ziway, Oromia region. Transmission intensity of *P. falciparum* and *P. vivax* is low. Samples for the current study were collected in the low transmission season in June and July 2019. Clinical samples were collected from individuals presenting with febrile illness to Batu and Dembel Health Centers. Cross-sectional surveys were conducted in 3 rural kebele (the lowest administrative units in Ethiopia), Bochessa, Dodicha, and Golba, which are under Adami Tulu Jiddo Kombolcha district administration.

### Sample and data collection

For the clinical samples, patients with suspected malaria infection were invited to join the study and provide an additional blood sample for diagnosis. A brief questionnaire was completed including age, sex, and kebele of residence of the patient. For community samples, a convenience sampling strategy was applied. The study team visited the villages, approached households, and asked all household members who were present to provide a sample. 100-200 µL blood were collected by finger prick into EDTA tubes. Blood samples were stored on ice packs in Styrofoam boxes until bringing them to the lab each evening, where they were stored at -20°C. RDT positive individuals among the community samples were referred to their health center for further diagnosis and treatment.

### Diagnosis by microscopy, RDT, and qPCR

Samples were collected by finger prick into EDTA tubes. All samples were screened by RDT (AccessBio CareStart Pf(HRP2)/Pv(LDH) combo) upon collection, and by local microscopy. For microscopy, WHO protocols were followed. 100 fields were assessed before declaring a sample negative.

DNA extraction was done using the Macherey-Nagel NucleoMag kit according to manufacturer’s instructions, with the following modification (Box 1xy): As the kit is optimized for extraction from 200 uL blood, but DNA was extracted from only 100 uL of blood, the volume of all reagents was reduced by 50%. Thus, per kit 8x 96 samples could be extracted, further reducing the amount of materials required and cost per sample. As proposed by the manufacturer as option, after the ethanol wash-step, beads were air-dried for 15 minutes instead of using buffer MBL-4. Some of the volumes of buffers were slightly modified to be able to complete all steps with a 30-300 µL multichannel pipette (Supplementary File S1). In a recent side-by-side comparison, the extraction kit used has yielded significantly more DNA than a spin-column based kit [28].

qPCR was done in a total volume of 12 µL, including 4 µL DNA, corresponding to 4 µL blood. For *P. falciparum* qPCR the *varATS* assay was used. This assay targets a multicopy gene that is present in 10-20 copies per parasite [29]. *P. vivax* qPCR was done using the *cox1* assay. This assay targets a mitochondrial gene that is present in approximately 10 copies per parasite [30]. Due to a manufacturing problem with the *P. vivax* probe (low yield), only a random subset of samples was screened by qPCR (653/1021 asymptomatic and 718/899 clinical samples). Infection prevalence and test positivity rate was compared among three age groups of 0<5 years, 5<15 years, and ≥ 15 years, between males and females, and between kebele (community sampling only) using Pearson’s Chi-square test.

HRP2-based RDTs can also detect the HRP3 protein, though sensitivity is lower [31]. Deletions of the *hrp2* gene result in false-negative RDTs in low-medium density infections. Deletions of *hrp2* and *hrp3* result in negative HRP2-based RDTs irrespective of parasite density [32, 33]. *P. falciparum* positive samples were typed for *hrp2*/*3* deletions by droplet digital PCR (ddPCR) [34]. For ddPCR, samples were shipped to the University of Notre Dame.

## Results

### Mobile laboratory

The mobile DNA extraction and qPCR systems were established in a makeshift laboratory on the compound of Addis Ababa University in Ziway. It consisted of a basic shed with two simple tables, and thus is representative for many locations with no laboratory infrastructure. All equipment and consumables required for this study are commercially available and given in Table 1. All protocols were optimized to achieve high throughput, i.e. work in 96-well format for extraction and 48-well format for the qPCR, while maintaining a low weight of the instruments required. Most importantly, the need for low weight instruments precluded the use of a centrifuge as used for common spin-column DNA extraction protocols. Instead, a protocol based on magnetic beads was used.

**Table 1:**
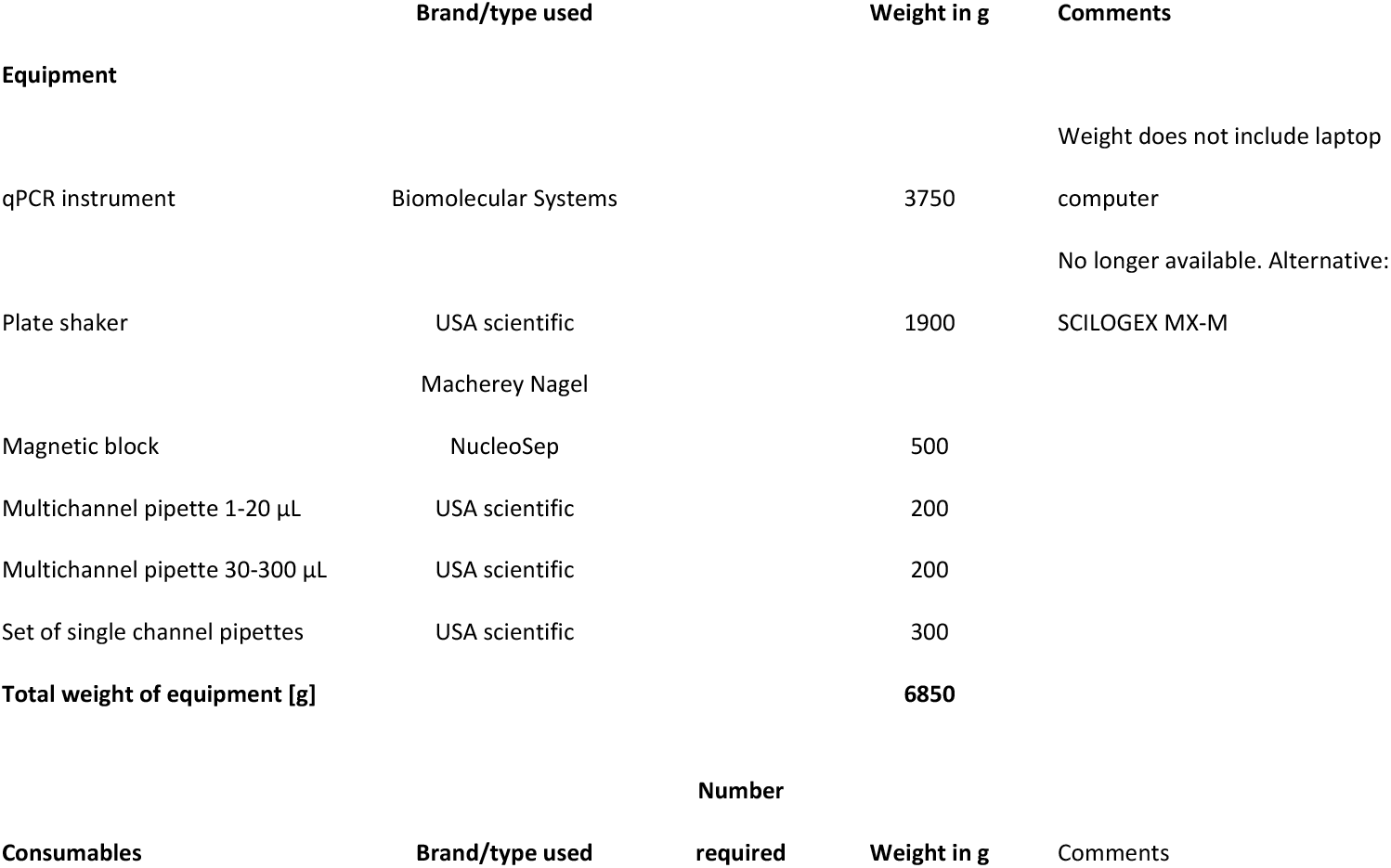
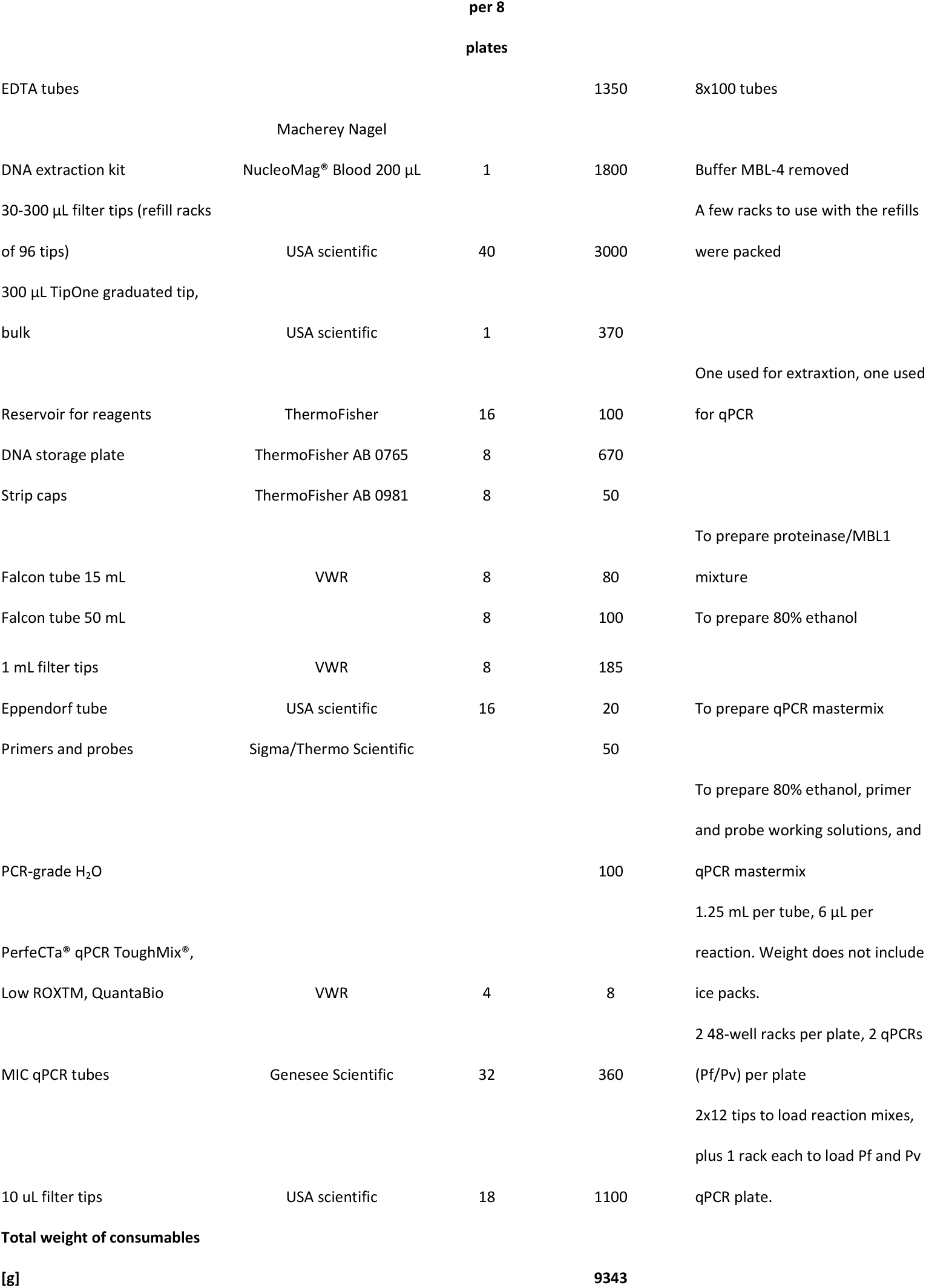
Equipment and consumables required for qPCR screening. One extraction kit lasts for 8 × 96-well plates. The the number and weight of all consumables and reagents reflects quantities required for 8 × 96 samples.

The main equipment required include the MIC qPCR system (including laptop computer), one plate shaker (for binding of DNA to the beads, wash steps, and DNA elution), one magnetic block (to bind the beads and remove the supernatant), two 12-channel pipettes (1-20 µL, 20-300 µL), and one set of single channel pipettes (1-20 µL, 20-200 µL, 100-1000 µL) (Figure 1). The total weight of all equipment was 6.9 kg, and it fit into one carry-on bag for air transport. The weight of all consumables for 8 plates (8×96 samples) was approximately 9.3 kg and thus can be easily transported by air as check-in luggage (Table 1). The cost of consumables including extraction kit, pipettes tips and other plasticware, and qPCR reagents was approximately USD 2.5 per sample.

**Figure 1:**
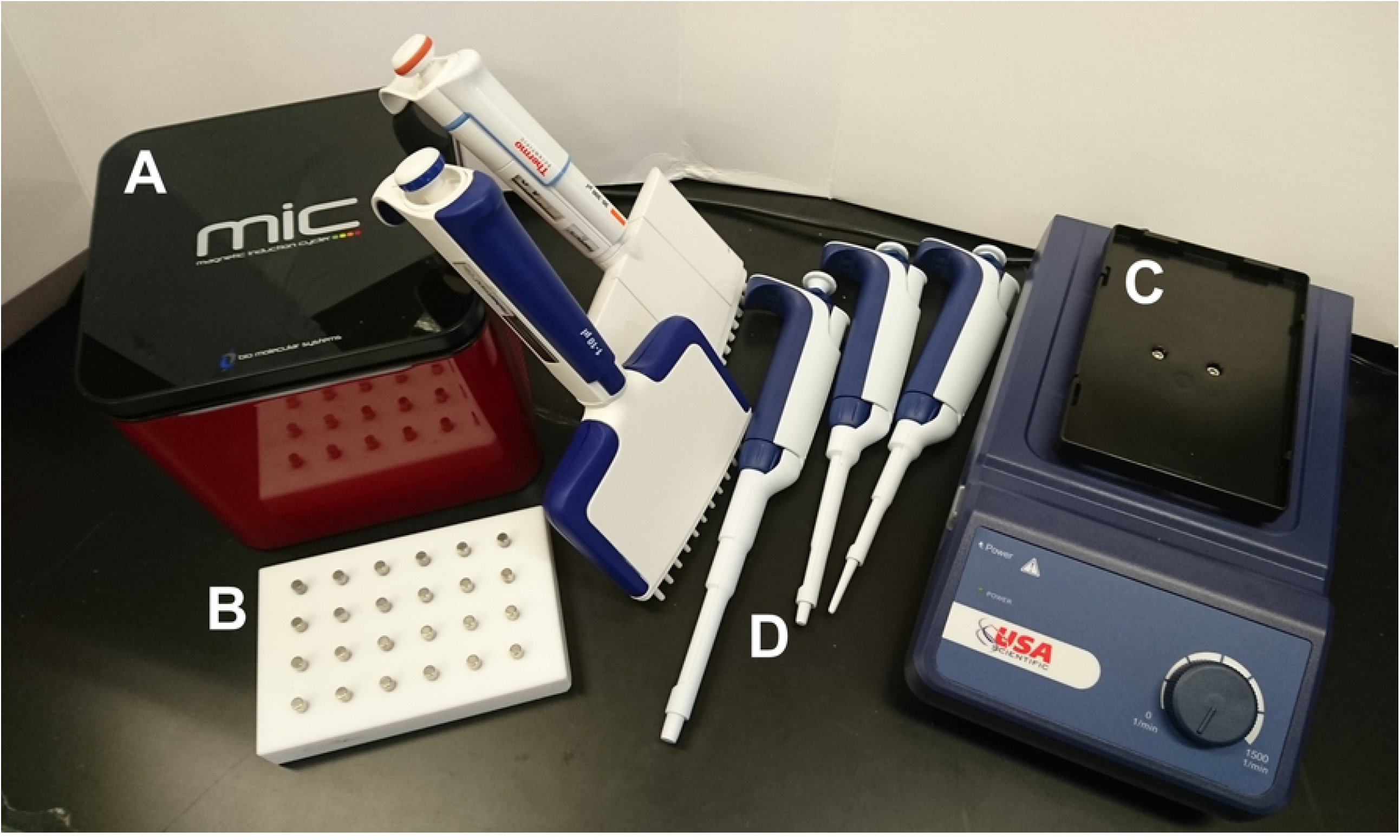
Instruments required for mobile qPCR. A) MIC 48-well qPCR instrument, B) magnetic block, C) plate shaker, D) pipettes.

The following items were purchased from local pharmacies: 99% Ethanol, nitrile gloves, lancets for blood sample collection, and microscopy slides. Further, a -20°C freezer was purchased locally for storage of reagents and samples. All extractions were done at room temperature. No water bath or incubator was needed.

### *P. falciparum* screening

Among 899 samples collected from febrile patients presenting to clinics, 51 (5.7%) were positive by qPCR. By local microscopy, only 13 were correctly diagnosed. One of the samples positive by qPCR for *P. falciparum* was misdiagnosed by microscopy as *P. vivax*. RDT was moderately more sensitive, with 18 samples positive for *P. falciparum*. A total of six samples were positive by microscopy and/or RDT, but not confirmed by qPCR (Figure 2). Demographic and qPCR data are given in Table 2. No significant difference in qPCR positivity by age group or sex was observed.

**Figure 2:**
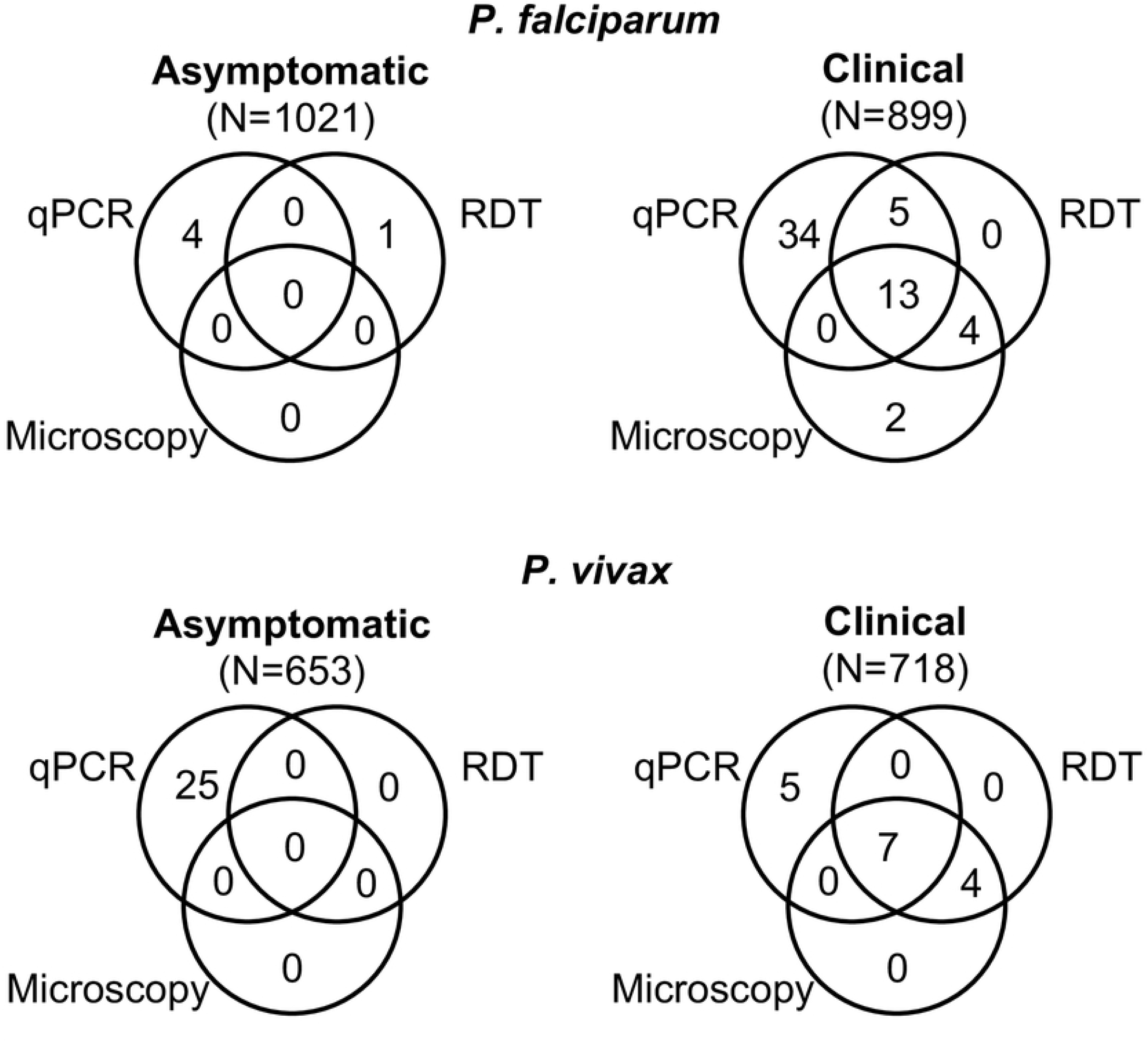
Number of samples positive by qPCR, RDT, and microscopy among clinical and asymptomatic individuals

**Table 2:**
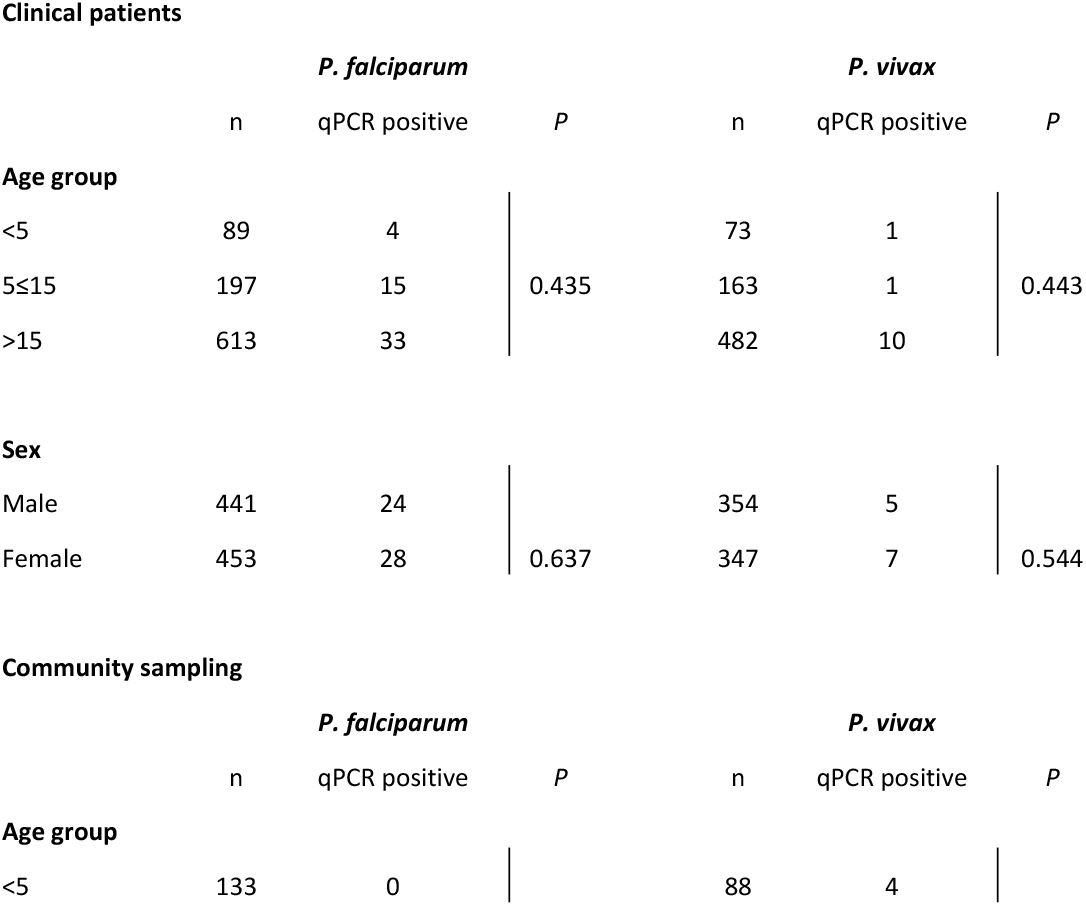
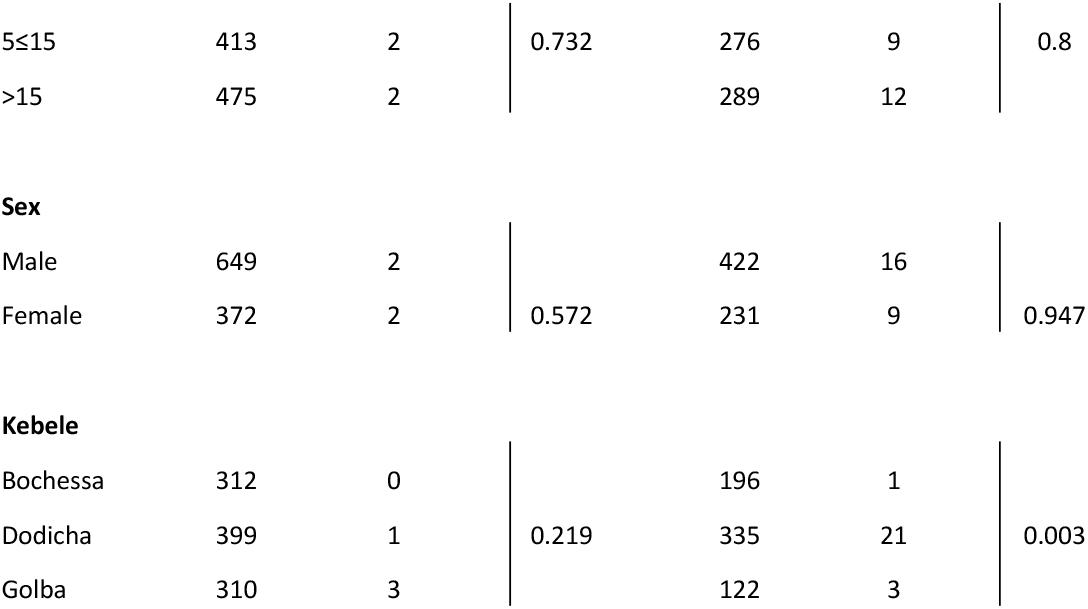
Demographic and qPCR data of study population

Clinical patients
|  | *P. falciparum* |  |  | *P. vivax* |  |  |
| --- | --- | --- | --- | --- | --- | --- |
|  | n | qPCR positive | *P* | n | qPCR positive | *P* |
| **Age group** |  |  |  |  |  |  |
| <5 | 89 | 4 | 0.435 | 73 | 1 | 0.443 |
| 5≤15 | 197 | 15 |  | 163 | 1 |  |
| >15 | 613 | 33 |  | 482 | 10 |  |
| **Sex** |  |  |  |  |  |  |
| Male | 441 | 24 | 0.637 | 354 | 5 | 0.544 |
| Female | 453 | 28 |  | 347 | 7 |  |

**Table 2:** Demographic and qPCR data of study population
|  | *P. falciparum* |  |  | *P. vivax* |  |  |
| --- | --- | --- | --- | --- | --- | --- |
|  | n | qPCR positive | *P* | n | qPCR positive | *P* |
| **Age group** |  |  |  |  |  |  |
| <5 | 133 | 0 |  | 88 | 4 |  |
| 5≤15 | 413 | 2 | 0.732 | 276 | 9 | 0.8 |
| >15 | 475 | 2 |  | 289 | 12 |  |
| **Sex** |  |  |  |  |  |  |
| Male | 649 | 2 |  | 422 | 16 |  |
| Female | 372 | 2 | 0.572 | 231 | 9 | 0.947 |
| **Kebele** |  |  |  |  |  |  |
| Bochessa | 312 | 0 |  | 196 | 1 |  |
| Dodicha | 399 | 1 | 0.219 | 335 | 21 | 0.003 |
| Golba | 310 | 3 |  | 122 | 3 |  |

*P. falciparum* prevalence among asymptomatic individuals was very low with only 4/1029 (0.4%) individuals positive by qPCR. None of them were positive by microscopy or RDT. One individual tested positive by RDT, but the infection was not confirmed by qPCR. The four qPCR positive individuals were 4, 14, 15, and 40 years old; two were male and two were female. Three of those positive were from Golba, and one from Dodicha (Table 2).

25 *P. falciparum* positive samples were successfully typed for *hrp2* deletion, and 24 samples for *hrp3* deletion. No *hrp2* deletions were observed, but 19/24 samples lacked the *hrp3* gene.

### *P. vivax* screening

Among 718 samples collected from febrile patients presenting to clinics and screened for *P. vivax* by qPCR, 12 (1.7%) were positive (Figure 2). Seven out of these twelve samples were also positive by microscopy and RDT. The twelve individuals that were positive by qPCR were 3 to 27 years old; with 10/12 being 15 years and older (Table 2). Four more samples were positive by microscopy and RDT, but not confirmed by qPCR. They remained negative when the qPCR was repeated. Mix up at the health center during sample and data collection might have occurred.

*P. vivax* prevalence among asymptomatic individuals was 3.8% (25/653). None of them was positive by microscopy or RDT (Figure 2). The age of positive individuals ranged from 1.5 to 60 years. No significant difference in prevalence rate among age groups was observed (Table 2). Prevalence differed significantly among kebele (*P*=0.003). It was highest in Dodicha at 6.3% (21/335), and lower in Golba at 2.5% (3/122) and Bochessa at 0.5% (1/196) (Table 2).

## Discussion

Molecular screening for infections below the limit of detection of microscopy or RDT is a key component of molecular malaria surveillance, and often the first step for subsequent studies, such as parasite genotyping to quantify drug resistance or establish transmission networks. Lack of adequate laboratory infrastructure is a problem in many endemic countries. In this study, using a mobile qPCR setup, high quality data was obtained from almost 2000 samples within a period of a few weeks. A short turnaround time is required in order to integrate molecular surveillance into control activities, for example to determine the extent of the asymptomatic reservoir during an outbreak [35].

The protocol tested is fully based on commercially available instruments and reagents, and offers high throughput with DNA extraction done in 96-well format, and qPCR run in 48-well format. A pair of trained laboratory technicians can process two 96-well plates within a day, and thus screen approximately 180 samples plus controls. The cost of all consumables for DNA extraction, and separate *P. falciparum* and *P. vivax* qPCR, is approximately 2.5 USD per sample. The protocol used is suited for extraction of any DNA or RNA from blood and thus can be applied for molecular diagnosis of any blood-borne pathogen. The protocol requires multiple pipetting steps and thus molecular laboratory skills are needed. Likewise, knowledge is required to interpret qPCR data. Training of malaria control program personnel will be crucial in order to integrate qPCR data into routine surveillance activities.

The makeshift laboratory presented multiple challenges. No running water was available, and power supply was unreliable with regular power cuts lasting several hours. As a result, the PCR was often run in the hotel, which had a back-up generator. For future surveillance by control programs, use of a generator to power the mobile lab is recommended. Of note, the DNA extraction can be done without the plate shaker, thus not requiring any power. Mixing steps can be done by pipetting. This protocol requires substantially more tips. The challenges of the mobile

lab were offset by the rapid availability of data. This was highlighted by the extended period of time required to obtain permit to ship samples to the US for *hrp2* and *hrp3* deletion typing.

This study revealed crucial reservoirs for transmission not identified by current control. Two thirds (34/52) of *P. falciparum* infections detected by qPCR in febrile patients were missed by microscopy and RDT. These untreated infections likely contribute to transmission for an extended period of time [36]. More sensitive diagnostic tools at health centers would be expected to reduce transmission. In contrast, very few infections were detected among asymptomatic individuals. A contrasting pattern was observed for *P. vivax*. Most infections were detected among asymptomatic individuals, and fewer among febrile patients. Asymptomatic *P. vivax* infections clustered mostly on one kebele. Possibly, many of the asymptomatic infections were relapses. Yet, while asymptomatic, they can still contribute to transmission [37]. This study corroborated high rates of subpatent *P. falciparum* and *P. vivax* infections in Ethiopia [38, 39]. Of note, transmission intensity in the Ziway region has declined drastically since 2005/2006, when a prevalence by microscopy of 16-19% was recorded [40].

In conclusion, this proof of concept study showed that actionable data on subpatent *P. falciparum* and *P. vivax* infections can be obtained in a short period of time using a mobile qPCR lab. Molecular screening for has identified a gap in the sensitivity for diagnosis of clinical *P. falciparum* cases and a substantial asymptomatic *P. vivax* reservoir, which was mostly concentrated in one village. *P. falciparum* control should focus on more sensitive diagnosis in health centers, e.g. though the introduction of novel, ultra-sensitive rapid diagnostic tests [41]. Prevention of onward transmission from the asymptomatic *P. vivax* reservoir might be achieved through improved vector control.

## Data Availability

All data is included in the manuscript and Supplementary File S2.

## Acknowledgements

We thank the study participants, kebele leaders, staff of AAU at Ziway satellite campus, staff at Batu and Dembel Health Centers, and the field collection teams.

## Supporting information caption

Supplementary File S1: DNA extraction protocol

Supplementary File S2: Database

## Notes

### Competing Interest Statement

The authors have declared no competing interest.

### Funding Statement

This work was supported by NIH R21AI137891 (CK). The funders had no role in study design, data collection and analysis, decision to publish, or preparation of the manuscript.

### Author Declarations

Informed written consent was obtained prior to sample collection from each study participant or, in the case of minors, from their parent or legal guardian. The study protocol was approved by the University of Notre Dame IRB (#19-03-5201), Trinity College Dublin, Addis Ababa University, and the Ethiopian National IRB.

## References

1. World Health Organisation. World malaria report 2021. 2021 978-92-4-004049-6.

2. Challi S, Miecha H, Damtie D, Shumie G, Chali W, Hailu T, et al. The Unmet Need: Low Performance of Laboratory Professionals in Malaria Microscopy, Oromia Regional State, Ethiopia. Am J Trop Med Hyg. 2020;102(1):117–20. Epub 2019/11/17. doi: 10.4269/ajtmh.19-0106. PubMed PMID: 31733053; PubMed Central PMCID: PMCPMC6947793.

3. Coleman RE, Maneechai N, Rachaphaew N, Kumpitak C, Miller RS, Soyseng V, et al. Comparison of field and expert laboratory microscopy for active surveillance for asymptomatic Plasmodium falciparum and Plasmodium vivax in western Thailand. Am J Trop Med Hyg. 2002;67(2):141–4. Epub 2002/10/23. doi: 10.4269/ajtmh.2002.67.141. PubMed PMID: 12389937.

4. Stuck L, Fakih BS, Al-Mafazy AH, Hofmann NE, Holzschuh A, Grossenbacher B, et al. Malaria infection prevalence and sensitivity of reactive case detection in Zanzibar. Int J Infect Dis. 2020;97:337–46. Epub 2020/06/14. doi: 10.1016/j.ijid.2020.06.017. PubMed PMID: 32534138.

5. Gamboa D, Ho MF, Bendezu J, Torres K, Chiodini PL, Barnwell JW, et al. A large proportion of P. falciparum isolates in the Amazon region of Peru lack pfhrp2 and pfhrp3: implications for malaria rapid diagnostic tests. PLoS One. 2010;5(1):e8091. Epub 2010/01/30. doi: 10.1371/journal.pone.0008091. PubMed PMID: 20111602; PubMed Central PMCID: PMCPMC2810332.

6. Gatton ML, Ciketic S, Barnwell JW, Cheng Q, Chiodini PL, Incardona S, et al. An assessment of false positive rates for malaria rapid diagnostic tests caused by non-Plasmodium infectious agents and immunological factors. PLoS One. 2018;13(5):e0197395. Epub 2018/05/15. doi: 10.1371/journal.pone.0197395. PubMed PMID: 29758050; PubMed Central PMCID: PMCPMC5951549.

7. Wongsrichanalai C, Barcus MJ, Muth S, Sutamihardja A, Wernsdorfer WH. A review of malaria diagnostic tools: microscopy and rapid diagnostic test (RDT). Am J Trop Med Hyg. 2007;77(6 Suppl):119–27. Epub 2008/01/31. PubMed PMID: 18165483.

8. Okell LC, Bousema T, Griffin JT, Ouedraogo AL, Ghani AC, Drakeley CJ. Factors determining the occurrence of submicroscopic malaria infections and their relevance for control. Nat Commun. 2012;3:1237. Epub 2012/12/06. doi: 10.1038/ncomms2241. PubMed PMID: 23212366; PubMed Central PMCID: PMCPMC3535331.

9. Cheng Q, Cunningham J, Gatton ML. Systematic review of sub-microscopic P. vivax infections: prevalence and determining factors. PLoS Negl Trop Dis. 2015;9(1):e3413. Epub 2015/01/09. doi: 10.1371/journal.pntd.0003413. PubMed PMID: 25569135; PubMed Central PMCID: PMCPMC4288718.

10. Bousema T, Okell L, Felger I, Drakeley C. Asymptomatic malaria infections: detectability, transmissibility and public health relevance. Nat Rev Microbiol. 2014;12(12):833–40. doi: 10.1038/nrmicro3364. PubMed PMID: 25329408.

11. Koepfli C, Nguitragool W, de Almeida ACG, Kuehn A, Waltmann A, Kattenberg E, et al. Identification of the asymptomatic Plasmodium falciparum and Plasmodium vivax gametocyte reservoir under different transmission intensities. PLoS Negl Trop Dis. 2021;15(8):e0009672. Epub 2021/08/28. doi: 10.1371/journal.pntd.0009672. PubMed PMID: 34449764; PubMed Central PMCID: PMCPMC8428688.

12. Tadesse FG, Slater HC, Chali W, Teelen K, Lanke K, Belachew M, et al. The Relative Contribution of Symptomatic and Asymptomatic Plasmodium vivax and Plasmodium falciparum Infections to the Infectious Reservoir in a Low-Endemic Setting in Ethiopia. Clin Infect Dis. 2018;66(12):1883–91. Epub 2018/01/06. doi: 10.1093/cid/cix1123. PubMed PMID: 29304258.

13. Almeida GG, Costa PAC, Araujo MDS, Gomes GR, Carvalho AF, Figueiredo MM, et al. Asymptomatic Plasmodium vivax malaria in the Brazilian Amazon: Submicroscopic parasitemic blood infects Nyssorhynchus darlingi. PLoS Negl Trop Dis. 2021;15(10):e0009077. Epub 2021/10/30. doi: 10.1371/journal.pntd.0009077. PubMed PMID: 34714821; PubMed Central PMCID: PMCPMC8555776.

14. Ouedraogo AL, Goncalves BP, Gneme A, Wenger EA, Guelbeogo MW, Ouedraogo A, et al. Dynamics of the Human Infectious Reservoir for Malaria Determined by Mosquito Feeding Assays and Ultrasensitive Malaria Diagnosis in Burkina Faso. J Infect Dis. 2016;213(1):90–9. Epub 2015/07/05. doi: 10.1093/infdis/jiv370. PubMed PMID: 26142435.

15. Sumner KM, Freedman E, Abel L, Obala A, Pence BW, Wesolowski A, et al. Genotyping cognate Plasmodium falciparum in humans and mosquitoes to estimate onward transmission of asymptomatic infections. Nat Commun. 2021;12(1):909. Epub 2021/02/12. doi: 10.1038/s41467-021-21269-2. PubMed PMID: 33568678; PubMed Central PMCID: PMCPMC7875998.

16. Faust CL, Brunker K, Ajambo D, Ryan M, Moses A, Rowel C, et al. Harnessing technology and portability to conduct molecular epidemiology of endemic pathogens in resource-limited settings. Trans R Soc Trop Med Hyg. 2020. Epub 2020/09/19. doi: 10.1093/trstmh/traa086. PubMed PMID: 32945867.

17. Inglis TJ, Bradbury RS, McInnes RL, Frances SP, Merritt AJ, Levy A, et al. Deployable Molecular Detection of Arboviruses in the Australian Outback. Am J Trop Med Hyg. 2016;95(3):633–8. Epub 2016/07/13. doi: 10.4269/ajtmh.15-0878. PubMed PMID: 27402516; PubMed Central PMCID: PMCPMC5014271.

18. Marx V. PCR heads into the field. Nat Methods. 2015;12(5):393–7. doi: DOI 10.1038/nmeth.3369. PubMed PMID: WOS:000353645800006.

19. Taylor BJ, Howell A, Martin KA, Manage DP, Gordy W, Campbell SD, et al. A lab-on-chip for malaria diagnosis and surveillance. Malar J. 2014;13:179. Epub 2014/06/03. doi: 10.1186/1475-2875-13-179. PubMed PMID: 24885206; PubMed Central PMCID: PMCPMC4029813.

20. Rampazzo RCP, Graziani AC, Leite KK, Surdi JA, Biondo CA, Costa MLN, et al. Proof of Concept for a Portable Platform for Molecular Diagnosis of Tropical Diseases: On-Chip Ready-to-Use Real-Time Quantitative PCR for Detection of Trypanosoma cruzi or Plasmodium spp. J Mol Diagn. 2019;21(5):839–51. Epub 2019/06/08. doi: 10.1016/j.jmoldx.2019.04.008. PubMed PMID: 31173930.

21. Take your lab wherever you go. The mobile genomics setup. Combines centrifuge, PCR and gel visualisation. Portable and ready-to-go. [cited 2021 January 04]. Available from: https://www.bento.bio.

22. miniPCR mini8 thermal cycler. Portable, durable 8-well PCR machine [cited 2021 January 04]. Available from: https://www.minipcr.com/products/minipcr/.

23. Biomeme. Real-time PCR anywhere you need it. [cited 2021 January 04]. Available from: https://biomeme.com.

24. Biomeme. Mobile qPCR Thermocyclers 2021 [cited 2021 November 14]. Available from: https://info.biomeme.com/mobile-qpcr-thermocyclers.

25. Taffese HS, Hemming-Schroeder E, Koepfli C, Tesfaye G, Lee MC, Kazura J, et al. Malaria epidemiology and interventions in Ethiopia from 2001 to 2016. Infect Dis Poverty. 2018;7(1):103. Epub 2018/11/06. doi: 10.1186/s40249-018-0487-3. PubMed PMID: 30392470; PubMed Central PMCID: PMCPMC6217769.

26. World Health Organisation. World Malaria Report 2020. 2020.

27. Wakabi W. Extension workers drive Ethiopia’s primary health care. Lancet. 2008;372(9642):880. Epub 2008/09/17. doi: 10.1016/s0140-6736(08)61381-1. PubMed PMID: 18795419.

28. Holzschuh A, Koepfli C. Tenfold difference in DNA recovery rate: systematic comparison of whole blood vs. dried blood spot sample collection for malaria molecular surveillance. Malar J. 2022;21(1):88. Epub 2022/03/17. doi: 10.1186/s12936-022-04122-9. PubMed PMID: 35292038.

29. Hofmann N, Mwingira F, Shekalaghe S, Robinson LJ, Mueller I, Felger I. Ultra-sensitive detection of Plasmodium falciparum by amplification of multi-copy subtelomeric targets. PLoS Med. 2015;12(3):e1001788. Epub 2015/03/04. doi: 10.1371/journal.pmed.1001788. PubMed PMID: 25734259; PubMed Central PMCID: PMCPMC4348198.

30. Gruenberg M, Moniz CA, Hofmann NE, Wampfler R, Koepfli C, Mueller I, et al. Plasmodium vivax molecular diagnostics in community surveys: pitfalls and solutions. Malar J. 2018;17(1):55. Epub 2018/01/31. doi: 10.1186/s12936-018-2201-0. PubMed PMID: 29378609; PubMed Central PMCID: PMCPMC5789620.

31. Kong A, Wilson SA, Ah Y, Nace D, Rogier E, Aidoo M. HRP2 and HRP3 cross-reactivity and implications for HRP2-based RDT use in regions with Plasmodium falciparum hrp2 gene deletions. Malaria J. 2021;20(1). doi: ARTN 207 10.1186/s12936-021-03739-6. PubMed PMID: WOS:000651405700003.

32. Cheng Q, Gatton ML, Barnwell J, Chiodini P, McCarthy J, Bell D, et al. Plasmodium falciparum parasites lacking histidine-rich protein 2 and 3: a review and recommendations for accurate reporting. Malar J. 2014;13:283. Epub 2014/07/24. doi: 10.1186/1475-2875-13-283. PubMed PMID: 25052298; PubMed Central PMCID: PMCPMC4115471.

33. World Health Organisation. P. falciparum hrp2/3 gene deletions. Conclusions and recommendations of a Technical Consultation. 2016.

34. Vera-Arias CA, Holzschuh A, Oduma CO, Badu K, Abdul-Hakim M, Yukich J, et al. Plasmodium falciparum hrp2 and hrp3 gene deletion status in Africa and South America by highly sensitive and specific digital PCR. 2021. doi: 10.1101/2021.06.01.21258117.

35. Recht J, Siqueira AM, Monteiro WM, Herrera SM, Herrera S, Lacerda MVG. Malaria in Brazil, Colombia, Peru and Venezuela: current challenges in malaria control and elimination. Malar J. 2017;16(1):273. Epub 2017/07/06. doi: 10.1186/s12936-017-1925-6. PubMed PMID: 28676055; PubMed Central PMCID: PMCPMC5496604.

36. Eichner M, Diebner HH, Molineaux L, Collins WE, Jeffery GM, Dietz K. Genesis, sequestration and survival of Plasmodium falciparum gametocytes: parameter estimates from fitting a model to malariatherapy data. Trans R Soc Trop Med Hyg. 2001;95(5):497–501. Epub 2001/11/15. doi: 10.1016/s0035-9203(01)90016-1. PubMed PMID: 11706658.

37. White MT, Karl S, Koepfli C, Longley RJ, Hofmann NE, Wampfler R, et al. Plasmodium vivax and Plasmodium falciparum infection dynamics: re-infections, recrudescences and relapses. Malar J. 2018;17(1):170. Epub 2018/04/19. doi: 10.1186/s12936-018-2318-1. PubMed PMID: 29665803; PubMed Central PMCID: PMCPMC5905131.

38. Assefa A, Ahmed AA, Deressa W, Wilson GG, Kebede A, Mohammed H, et al. Assessment of subpatent Plasmodium infection in northwestern Ethiopia. Malar J. 2020;19(1):108. Epub 2020/03/07. doi: 10.1186/s12936-020-03177-w. PubMed PMID: 32131841; PubMed Central PMCID: PMCPMC7057598.

39. Tadesse FG, Pett H, Baidjoe A, Lanke K, Grignard L, Sutherland C, et al. Submicroscopic carriage of Plasmodium falciparum and Plasmodium vivax in a low endemic area in Ethiopia where no parasitaemia was detected by microscopy or rapid diagnostic test. Malar J. 2015;14:303. Epub 2015/08/06. doi: 10.1186/s12936-015-0821-1. PubMed PMID: 26242243; PubMed Central PMCID: PMCPMC4524028.

40. Kibret S, Alemu Y, Boelee E, Tekie H, Alemu D, Petros B. The impact of a small-scale irrigation scheme on malaria transmission in Ziway area, Central Ethiopia. Trop Med Int Health. 2010;15(1):41–50. Epub 2009/11/18. doi: 10.1111/j.1365-3156.2009.02423.x. PubMed PMID: 19917039.

41. Danwang C, Kirakoya-Samadoulougou F, Samadoulougou S. Assessing field performance of ultrasensitive rapid diagnostic tests for malaria: a systematic review and meta-analysis. Malar J. 2021;20(1):245. Epub 2021/06/05. doi: 10.1186/s12936-021-03783-2. PubMed PMID: 34082776; PubMed Central PMCID: PMCPMC8176703.

